# Return To Sporting Activity After Total Hip Arthroplasty - An Irish Experience

**DOI:** 10.1101/2020.04.24.20077784

**Authors:** Darren Patrick Moloney, Danilo Vukanic, Ronan Kearney, Marc C Grant-Freemantle, John F Quinlan

## Abstract

**Objective:** Total hip arthroplasty (THA) is traditionally an operation of the elderly. In the past few decades we have seen younger patient cohorts undergoing THA and increased activity levels in elderly cohorts. Guidelines have not been developed on return to sporting activity after THA. At present return to sport is dictated by surgeon preference and advice. The aim of this paper is to assess attitudes and practices toward return to sport after THA in Irish surgeons performing a minimum of forty total hip replacements per annum.

**Design:** Web-based survey, level of evidence 5

**Methods:** Current practices in the Irish orthopaedic community were assessed through a web-based survey. The questionnaire was issued via the Royal College of Surgeons Ireland to consultants involved with higher specialist training. Of 90 consultants surveyed, 35 responded. 8 respondents did not perform greater than forty THA operations per annum and thus were excluded from the later part of the survey.

**Results:** The majority of respondents (61.54%) would use an uncemented design for a patient planning on returning to sport. 26.92% would use a hybrid design. 3.85% would opt for a resurfacing arthroplasty. The majority of surgeons (73.08%) would use an uncemented femoral implant. 92.81% would use a ceramic on polyethylene bearing surface. 57.69% of surgeons would perform a posterolateral approach and 30.77% would opt for a anterolateral approach. All respondents would allow patients to return to low impact sports such as golf, swimming and walking following THA. Significantly more variance was seen across surgeons when considering a return to medium impact and high impact sports.

**Summary/Conclusion:** This survey has shown that there is still a lack of consensus on return to sport after THA. This survey of some of the most eminent arthroplasty surgeons practicing in Ireland will hopefully allow for consolidation of guidelines on return to sport after THA.

## INTRODUCTION

Hailed as the “operation of the century”, total hip arthroplasty (THA) is one of the most ubiquitous and successful operations in orthopaedic practice today.[1] Pioneered by the father of arthroplasty, Sir John Charnley in the 1960s, THA has become universal. THA was the 26th most commonly performed procedure in the United States in 2015, with an estimated 300’000 cases per annum.[2] In 2014, approximately 40% of THA was performed on patients younger than 65 years of age in the United States.[2]

With the numbers of THA increasing every year there is inevitably an increase in younger more active patients undergoing this procedure.[3] Crownshield et al. predicted an increasingly younger and more active cohort undergoing joint replacement surgery.[4] The Irish National Orthopaedic Registry is currently in the process of establishing itself as the national registry for arthroplasty, however, it has not yet incorporated all Irish hospitals.[5] It is therefore unclear at present the exact number of patients undergoing THA each year in Ireland. In 2005, Curtin et al. surveyed Irish arthroplasty surgeons and identified that practices varied when performing THA in younger cohorts.[6] 69% of surgeons surveryed noted to use a different THA in younger patient cohorts when compared with elderly cohorts. In addition, 70% of surgeons change implant design and 47% use different bearing surfaces for the different age cohorts.

The evolving demographics in the practice of arthroplasty with younger cohorts undergoing joint replacement surgery requires consensus on return to sport after such surgery. Bradley et al. surveyed members of the British Hip Society to ascertain return to athletic activity recommendations after total hip replacement of British hip surgeons. Our study aims to assess attitudes and practices toward return to sport after THA in Irish hip surgeons.

### DESIGN

#### Data Collection

Irish orthopaedic surgeons were identified through communication with the Royal College of Surgeons Training Committee. The web-based survey was designed using surveymokey.com and was distributed throughout to the consultants by email.

Following participant consent, Consultants were questioned on their experience in terms of number of years working as well as the number of THA they perform per annum. Participants answered further questions on returning to low, medium and high impact sports following THA, similar to Clifford et al..[7]

Surgeons were also asked questions based on their preferred surgical technique for patients undergoing THA who wished to return to sport.

#### Inclusion Criteria

Consultant Orthopaedic surgeon practicing in Ireland and registered with the Irish Surgical Training Programme.

#### Exclusion Criteria

The design of the web-based survey eliminated surgeons who performed less than forty total hip replacements per annum. They were as a result not asked to give feedback on the later part of the survey where attitudes toward returning to sport after total hip arthroplasty were assessed.

#### Questionnaire Design

The questionnaire was designed in six sections. The first section focussed on surgeon demographics. It assessed the number of hip replacements they performed per annum, the number of years they had been practicing as a surgeon and whether the surgeon considered themselves a specialist in arthroplasty of the young.

The next section asked when would you allow a patient to return to sports after total hip arthroplasty.

The following three sections explored whether a surgeon would allow or allow with experience, i.e. a graduated physiotherapy programme or not allow a patient to return to sports. The sections covered 22 different sports and classified them as low, medium and high impact.

The final section examined implant choice for a patient wishing to return to sport. The surgeons were questioned on their overall design choice, their femoral implant choice along with their bearing surface and head size choice. Finally the surgeons were questioned on their preferred surgical approach in such cases.

## RESULTS

90 Irish Orthopaedic consultants were contacted as part of this study. 35 consultants responded. The response rate for the survey was 38.89%. Of these surgeons 8 did not perform greater than 40 total hip replacements per annum and were therefore excluded from the latter half of the survey. 40% of respondents performed over 100 THAs per annum. 77.14% performed greater than 40 THA per annum. 91.43% of surgeons had been performing for greater than 5 years as a consultant arthroplasty surgeon. 40% considered themselves specialists in hip replacement in the young active patient and overall 74.29% would be happy to operate on a patient of any age. This is illustrated in table 1.

**Table 1:** Proportion of Surgeons With Special Interest in Young Adult Total Hip Arthroplasty

| Special Interest in Young Adult THA | Proportion of Irish High Volume Arthroplasty Surgeons: Responses |
| --- | --- |
| Yes | 40% (14) |
| Not particularly but happy to operate on any age | 34.29% (12) |
| No | 25.71% (9) |

Surgeon responses for recommendations of returning to various sports stratified by impact level are tabulated in table 2. All surgeons would allow return to low impact sporting activities such as golf, walking, swimming, cross training and static cycling.

**Table 2:** High Volume Hip Arthroplasty Surgeons Responses for Return to Sporting Activity After Total Hip Arthroplasty (THA) Classified According to Impact Category

| Sport | Recommendation of high volume Irish hip arthroplasty surgeons |  |  |  |
| --- | --- | --- | --- | --- |
|  | Allow | Allow with Experience | Not Allow | Undecided |
| *Low Impact Sports* |  |  |  |  |
| Golf | 96.15% | 3.85% | 0.00% | 0.00% |
| Swimming | 92.31% | 7.69% | 0.00% | 0.00% |
| Walking | 96.15% | 3.85% | 0.00% | 0.00% |
| Cross Training | 80.77% | 15.38% | 0.00% | 3.85% |
| Static Cycling | 92.31% | 7.69% | 0.00% | 0.00% |
| *Medium Impact Sports* |  |  |  |  |
| Cycling | 92.31% | 7.69% | 0.00% | 0.00% |
| Hiking/Hill-walking | 92.31% | 3.85% | 3.85% | 0.00% |
| Dancing | 80.77% | 19.23% | 0.00% | 0.00% |
| Weightlifting (high weight/low reps) | 30.77% | 38.46% | 30.77% | 0.00% |
| Ice skating/Roller Blading | 30.77% | 42.31% | 23.08% | 3.85% |
| Rowing | 46.15% | 46.15% | 3.85% | 3.85% |
| Bowling | 80.77% | 19.23% | 0.00% | 0.00% |
| Low Impact Aerobics | 80.77% | 15.38% | 0.00% | 3.85% |
| *High Impact Sports* |  |  |  |  |
| Contact sports | 7.69% | 34.62% | 57.69% | 0.00% |
| Jogging (Treadmill) | 57.69% | 19.23% | 23.08% | 0.00% |
| Jogging (Road) | 38.46% | 19.23% | 42.31% | 0.00% |
| Squash | 26.92% | 19.23% | 53.85% | 0.00% |
| Martial Arts | 3.85% | 23.08% | 69.23% | 3.85% |
| Cricket | 50.00% | 26.92% | 15.38% | 7.69% |
| High Impact Aerobics | 30.77% | 23.08% | 42.31% | 3.85% |

Medium impact sports were generally allowed or allowed with experience. One surgeon would not recommend returning to hiking post THA. Interestingly the exception in medium impact sporting activities was weightlifting and ice-skating. 30.77% would not allow patients to return to weightlifting (high weight/low repetition) weightlifting. Ice-skating/ rollerblading was also not recommended by 1 in 4 of the respondents (23.08%). A single surgeon in the group would not recommend returning to rowing post THA. High impact sports were not recommended by a large proportion of Irish arthroplasty surgeons. The majority 57.69% would not recommend return to contact sports after THA. Timing of return to sport after THA was varied. 7.41% would recommend immediate return to sport. 18.52% would recommend return to sport within 6-12 weeks. The majority (59.26%) recommended return to sport between 3 and 6 months. This is illustrated in table 3.

**Table 3:**
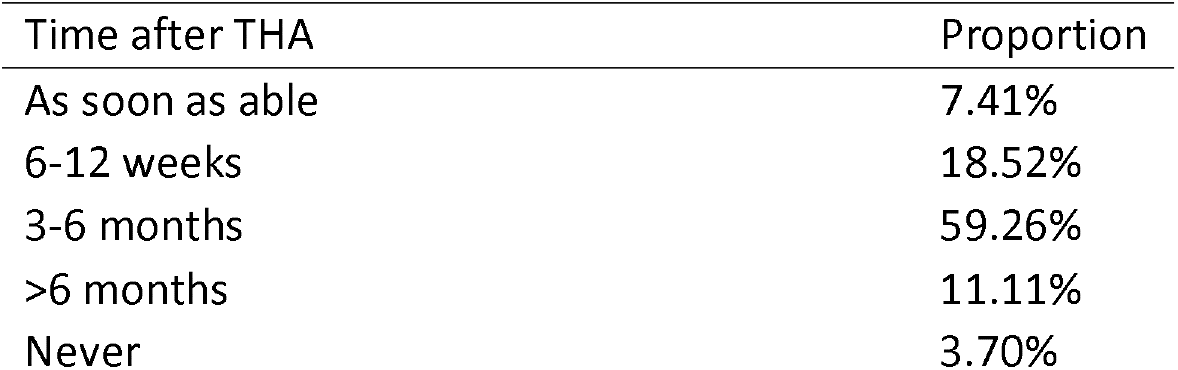
Time Interval Recommended Before Allowing Return to Sporting Activities After Total Hip Arthroplasty (THA)

| Time after THA | Proportion |
| --- | --- |
| As soon as able | 7.41% |
| 6-12 weeks | 18.52% |
| 3-6 months | 59.26% |
| >6 months | 11.11% |
| Never | 3.70% |

Implant choice, THA design and bearing couple was also evaluated as part of this study. The majority of Irish hip surgeons would perform an uncemented design (61.54%). 1 in 4 would perform a hybrid design 26.92%. Only one surgeon reported that they would opt for resurfacing arthroplasty in the patient who wishes to return to sport. These results are summarised in table 4.

**Table 4:** Preferred Implant Choice in Sporting Patient Undergoing Total Hip Arthroplasty (THA).

| Design | Proportion |
| --- | --- |
| Hybrid | 26.92% |
| Cemented | 7.69% |
| Uncemented | 61.54% |
| Resurfacing Arthroplasty | 3.85% |

In terms of the femoral stem 73.38% of Irish surgeons would perform uncemented femoral stems. Head size of less than 36mm was favoured by 76.92% of respondents. Ceramic-on-polyethylene was recommended by the majority of surgeons (92.31%). Bearing choice is summarised in table 5.

**Table 5:** Preferred Bearing Surface in Sporting Patient Undergoing Total Hip (THA). Arthroplasty

| Bearing Couple | Responses |
| --- | --- |
| Metal on Polyethylene | 3.85% |
| Ceramic on Ceramic | 3.85% |
| Ceramic on Polyethylene | 92.31% |
| Metal on metal | 0.00% |
| Other (please specify) | 0.00% |

The approach most commonly chosen in the setting of a young patient wishing to return to sport was the posterolateral approach 57.69%, followed by direct lateral 30.77%. It was unspecified whether trans-trochanteric or trans-gluteal was preferred in this case. 11.54% would opt for direct anterior approach. Preferred Surgical Approach is summarised in table 6.

**Table 6:** Preferred Surgical Approach in Sporting Patient Undergoing Total Hip Arthroplasty (THA)

| Surgical Approach | Responses |
| --- | --- |
| Posterior | 57.69% |
| Direct Anterior | 11.54% |
| Anterolateral | 30.77% |
| Other (please specify) | 0.00% |

## DISCUSSION

### Current Recommendations

Although many arthroplasty surgeons have anecdotes of young patients who returned to sport at a high level post-operatively, the plural of anecdote is not data. There is a lack of empirical data to suggest the appropriate timeframe for return to sports post total hip replacement. There are very few studies which evaluate the safety of various sports for individuals who have undergone THA. Most of the current evidence is consensus opinion i.e. level 5 evidence.[6, 8, 9] The current recommendations for return to sport after total hip arthroplasty are from surveys of hip and knee surgeons based on clinical experience and preference.

### International attitudes toward return to sport after THA

Klein et al. examined return to athletic activity after THA in a 2005 study, examining 614 American surgeons, from the Hip Society and the American Association of Hip and Knee Surgeons.[8] The surgeons were asked a set of similar questions to the ones we used in this study. In general, low impact sports were allowed post THA such as swimming, bowling and stationary biking. High impact sports such as squash, jogging and contact sports were not allowed post THA. For allowable activities the recommended timeframe was 3-6 months following surgery and approximately 1 in 3 surgeons would allow their patients to return to sport after 1-3 months.

Swanson et al. looked at group of 139 hip surgeons and asked them to categorise their recommendations for various activities.[10] They used an approach of asking whether a patient should be allowed to perform the activity “unlimited”, “occasional (1-2 times per month)” or “discouraged”. Low impact activities such as cycling, walking and golf were generally allowed with unlimited frequency. None of the surgeons indicated any scientific rationale for their recommendations.

Wylde et al. studied return to sport after joint replacement in 2008.[9] They followed patients three years after their index procedure. Of their 2085 patients undergoing various forms of arthroplasty, 726 (34.8%) performed sporting activities prior to their operation. 446 (61.4%) returned to sport post-operatively. The most common pre-operative sporting activities were swimming, walking and golf. Return to sport was not significantly different across THA and hip resurfacing.

Bradley et al. performed a similar study with British Hip Society surgeons.[11] 109 members were questioned in this study. 67.6% had an interest in hip replacement surgery in younger patients. 67.9% performed greater than 100 THAs per year. Again, all surgeons recommended low impact sports such as walking, swimming and golfing. Interestingly 44.8% would not recommend weight training post THA compared to 30.77% of Irish surgeons in our study. Interestingly 8.6% would not recommend rowing perhaps due to the extreme flexion associated with this activity. 43.7% would advise waiting three months after surgery to return to sport, by comparison our study found 59.26% of surgeons would advise return to sport after 3 months.

### Previous attitudes in Ireland

Curtin et al. performed a questionnaire based study of surgeons in Ireland in 2005.[6] 9% of surgeons at this point were unhappy to perform surgery on a younger patient and referred these patients to a colleague. 45% used a hybrid design, 15% used an uncemented THA and 15% performed hip resurfacing. By comparison our study showed that only 3.85% would recommend resurfacing arthroplasty. 17% of surgeons in Ireland in 2005 used Charnley prosthesis in the young. Our study did not take in to account specific manufacturer implant choice.

### Individual sports after THA

A number of studies suggest the consensus that low impact sports are generally recommended after THA. The low impact sports included walking, stationary cycling, dancing, golf, bowling and swimming.[9-11] The studies suggest that contact sports such as soccer, football and basketball are generally not recommended following THA. Other high impact sports such as running was also not recommended by the plurality in these studies..[11)

D’Lima et al. described contact forces in the operated leg during various activities.[12] Stationary cycling was noted to evoke contact stress at the knee joint 1.3 times greater than body weight (BW). In golf the leading leg is under 4.5 × BW and 3.2 × BW in the following leg. Whilst jogging the leg is under 3.6 × BW and walking 2.6x BW. However, this study pertains to joint reaction forces at the knee joint.

### THA Design in the Patient Wishing to Return to Sport

Bradley et al. reports on THA designs in the setting of a patient wishing to return to sport. The preferred implant choice was uncemented similar to our study. A minority say they would perform a completely cemented hip replacement. Interestingly 11.7% would perform hip resurfacing however only 3.85% in Ireland would perform hip resurfacing. This is perhaps due to Ireland’s chequered history with metal on metal hip replacements. Bearing choice in the British group was mostly ceramic on ceramic, however in the Irish group ceramic on polyethylene was chosen by the majority of surgeons which is in line with current evidence on the least wear in bearing surfaces.[13, 14]

## Data Availability

Data are available upon reasonable request. Data were sourced from online web based survey in a GDPR compliant manner. Consolidated deidentified participant data are available from Darren Moloney, ORCID identifier 0000-0002-6530-1467. There is no statistical analysis as part of this study.

## SUMMARY

### What are the new findings?

Irish Orthopaedic Surgeon attitudes have changed in the last 15 years regarding implant choice in the young patient undergoing THA. Fewer surgeons are performing hip resurfacing. Irish surgeons also appear to be less inclined to recommend hip resurfacing than other countries. Ceramic-on-polyethylene is the most recommended bearing couple choice in Ireland for patients wishing to return to sport.

### How might it impact on clinical practice in the future?

This study is further level 5 evidence pertaining to hip replacement in the young active patient. Further study is required to evaluate the risks and benefits of returning to various forms of sport after THA.

## INTELLECTUAL PROPERTY RIGHTS ASSIGNMENT OR LICENCE STATEMENT

I, Darren Moloney, the Author has the right to grant and does grant on behalf of all authors, an exclusive licence and/or a non-exclusive licence for contributions from authors who are: i) UK Crown employees; ii) where BMJ has agreed a CC-BY licence shall apply, and/or iii) in accordance with the relevant stated licence terms for US Federal Government Employees acting in the course of the their employment, on a worldwide basis to the BMJ Publishing Group Ltd (“BMJ”) and its licensees, to permit this Work (as defined in the below licence), if accepted, to be published in British Journal of Sports Medicine and any other BMJ products and to exploit all rights, as set out in our licence.

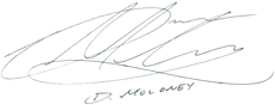

Date: 24th of April 2020

## COMPETING INTERESTS

No, there are no competing interests for any author

Date: 24th of April 2020

## CONTRIBUTORSHIP

DP Moloney, conception, writing, literature review

D Vukanic, literature review, proofing

R Kearney, conception, proofing, literature review

MC Grant-Freemantle, proofing

JF Quinlan, conception, organisation, proofing

Date: 24th of April 2020

## ACKNOWLEDGEMENTS

The authors do not wish to make any acknowledgements.

Date: 24th of April 2020

## DATA SHARING STATEMENT

Date: 24th of April 2020

## FUNDING INFO

The author(s) disclosed receipt of the following financial support for the research, authorship, and/or publication of this article: There was no funding received.

Date: 24th of April 2020

## ETHICAL APPROVAL INFORMATION

Ethics approval was sought for this publication from an institutional ethics committee. This paper describes an anonymous web based survey of consultant arthroplasty surgeons so did not meet the requirements for ethical review process.

Date: 10th of February 2020

